# Real-world evidence: telemedicine for complicated cases of urinary tract infection

**DOI:** 10.1101/2022.10.03.22280523

**Authors:** Natalie M. Daumeyer, Daniel Kreitzberg, Kathleen M. Gavin, Timothy A. Bauer

## Abstract

**Background:** Telemedicine programs for the treatment of urinary tract infections (UTIs) offer an opportunity to reduce burdens on patients and providers. However, these programs are typically restricted to patients with uncomplicated UTIs. This real-world analysis evaluated treatment and resolution rates in a large-scale, national UTI telemedicine program inclusive of patients with uncomplicated and complicated UTIs.

**Methods and findings:** We conducted a retrospective analysis of data obtained from a commercially available telemedicine program for the treatment of UTIs among adult women in the US between 2017 and 2021 (n=51,474). The primary outcomes were the number of women who presented with symptoms of uncomplicated UTI, complicated UTI, and vaginal infection; prescription use and antibiotic type; symptom resolution within 7 days after appointment; and treatment failure or relapse. Most patients reported frequent urination (94.4%), urgency (94.5%), and dysuria (97.6%). Those with uncomplicated UTI symptoms represented the majority of patients (61.6%); however, a substantial number of patients (36.5%) also reported at least 1 symptom associated with a complicated UTI. One-fifth of patients (19.2%) reported at least 1 co-occurring symptom of vaginal infection or sexually transmitted infection. Across all treated patients, 94.0% received recommended antibiotics according to the clinical protocol. Of the treated patients who provided follow-up data (n=3,521), 89.7% reported 7-day symptom resolution. Symptom resolution rates were similar between patients with uncomplicated UTI symptoms (90.8%) and complicated UTI symptoms (87.9%), and symptom resolution among all treated patients (89.7%) was similar to reports for in-person standard of care.

**Conclusions:** These findings suggest that large-scale telemedicine programs for the treatment of UTIs can be effective in the treatment of complicated UTIs.

## Introduction

Urinary tract infections (UTIs) are some of the most common infections, accounting for millions of visits to healthcare facilities every year [1]. The use of urinalysis in emergency departments to diagnose suspected UTIs has increased from 23% in 2007 to 27% in 2016 [2]. During these visits, patients can spend more than 2 hours waiting for and receiving care, creating a burden for patients [3]. Using telemedicine as an alternative model of care for patients with UTIs would reduce the amount of time patients spend seeking treatment, while freeing up important resources in often over-burdened urgent care and emergency departments, as well as primary care clinics [4,5]. UTIs make up 0.7% of outpatient services, according to projections from US registries [1]. Between 1996 and 2001, an average of 7 million women visited healthcare centers in the US for uncomplicated UTIs [6]. There were 10.5 million ambulatory visits for UTIs in the US in 2007 (accounting for approximately 1% of all ambulatory visits), of which, 21.3% were to emergency departments [7]. Consultations for UTIs represent between 1% and 6% of all medical visits (∼7□million visits and ∼$1.6□billion□annually) [8]. Previous research has shown that telemedicine treatment programs are effective for women presenting with uncomplicated UTI symptoms [9-15]. These programs, which largely rely on the application of inclusion/exclusion criteria according to self-reported symptoms and empirical use of antibiotics [16], are similar to in-person care and reduce time to recovery compared with diagnosis using urinalysis [16,17]. However, a limitation of these programs is the exclusion of patients who self-reported symptoms of complicated UTIs (e.g., fever, nausea) or vaginal infections (e.g., vaginal discharge, vaginal irritation) [9,11-14]. Thus, it is unclear how effective or safe telemedicine programs can be for patients with these clinical profiles.

Women with symptoms of complicated UTIs (e.g., pyelonephritis) are usually referred to in-person care to receive intravenous antibiotics and/or undergo urinalysis to determine the appropriate antibiotic regimen [18,19]. However, some complicated UTI cases can be managed at home without admittance to an emergency department. Researchers have thus begun to question whether higher-risk patients, such as those who report symptoms of pyelonephritis or more complicated UTI, can be treated via telemedicine [15,18,20].

In a similar manner, women with vaginal symptoms are often referred to in-person care to rule out other diagnoses, such as sexually transmitted infections (STIs) and vaginal infections [16,21,22]. However, even cases of suspected STI may be managed by remote sample collection and telemedicine services [23]. Recent evidence has shown that vaginal discharge does not reduce the likelihood of a UTI [24], contradicting earlier studies advocating for in-person treatment for women with vaginal symptoms [16]. Thus, with proper symptom screening, which can be implemented virtually, a process for ruling in or out vaginal infections and STIs may be accomplished [25]. Patients with symptoms of complicated UTI or vaginal infection may benefit from a telemedicine approach to care, but current studies evaluating the value of telemedicine have excluded these patients. It remains unknown whether telemedicine provides an alternative, effective pathway to care without compromising on quality.

This study examined real-world data from a large telemedicine program that sought to provide treatment for women with UTI symptoms. This UTI telemedicine program is different from previously evaluated programs [15] in the following ways: 1) symptoms reported during the initial screening are included, 2) patients who reported symptoms of complicated UTI and vaginal infection were included, and 3) physicians were given discretion as to whether and how to treat women who reported symptoms of complicated UTI or vaginal infection. The primary outcome measures were the number of women who presented with symptoms of uncomplicated UTI, complicated UTI, and vaginal infection; prescription use and antibiotic type; symptom resolution within 7 days after appointment; and treatment failure or relapse.

## Methods

### Ethics

This study is a retrospective analysis of real-world data from a nationwide commercial telemedicine program for the treatment of UTIs. The project was deemed exempt from IRB review by WCG Institutional Review Board (IRB00000533) because it does not meet the definition of human subjects research as defined in federal regulation 45 CFR 46.102.

### Study population and data collection

Individuals in the US seeking care for UTI symptoms between November 2017 and November 2021 completed a digital screening process to determine their eligibility for telemedicine services. Patients were excluded for the following reasons: younger than 18 years of age, male, pregnant or breastfeeding, immunocompromising conditions (e.g., HIV, lupus), history of kidney condition, history of urinary condition, history of urologic procedure, history of resistant organisms, or inpatient stay at a healthcare facility within the past 3 months. For this protocol, prior to the COVID-19 pandemic, women aged 65 years or older were excluded if they were currently taking more than 5 prescription medications; this contraindication was removed during the pandemic because of restricted access to in-person care. Cancelled/missed appointments, duplicate records, and follow-up appointments occurring within 30 days after the initial appointment were excluded from the analysis set. Those meeting the criteria reported their symptoms through the digital screener and scheduled a telemedicine consultation. Patients who reported symptoms of a complicated UTI or vaginal infection were able to schedule a telemedicine consultation. Immediately following their consultation, patients received a satisfaction questionnaire. After 7 days, patients received a follow-up questionnaire to assess their symptom resolution. All patients who reported unresolved symptoms received follow-up telemedicine services. Fig 1 shows the participant selection process for this cohort study.

**Fig 1.**
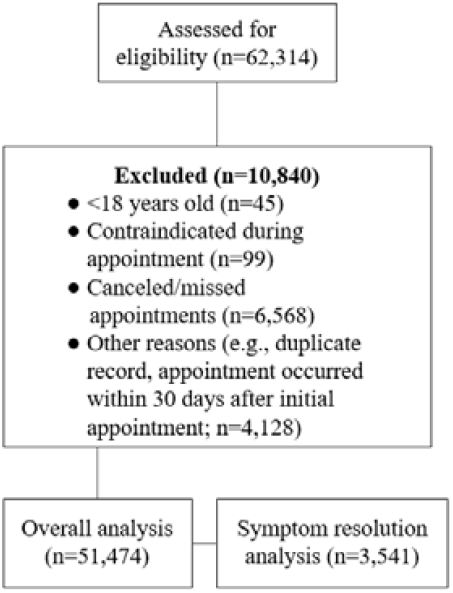
Study flow chart

### Physician information and the clinical protocol

The median number of consultations was 79 (IQR=623) among 84 physicians. Physician specialties varied and included 51.2% (n=43) family medicine, 29.8% (n=25) internal medicine, 11.9% (n=10) emergency medicine, and 4.8% (n=4) preventative medicine, as well as 1 physician who was trained in both internal and emergency medicine and 1 physician who was trained as a radiologist. A clinical protocol provided guidance for physicians to diagnose and treat uncomplicated UTIs using the telemedicine platform. The protocol outlined contraindications, considerations for treatment, and recommended antibiotic regimens (see S1 Appendix) [18,22,26]. All physicians were trained on the protocol, which was available for reference during consultations. The protocol recommended that patients with symptoms of complicated UTIs or vaginal infection be referred to in-person care. However, physicians were permitted to use their discretion to treat via telemedicine or refer to in-person care. STIs were ruled out during physician consultation based on answers provided on the questionnaire. If providers were unsure about treating a patient, they could escalate the issue to the telemedicine program directors for additional guidance.

### Measures

#### Demographic characteristics

Patients were asked their age (in years) and sex during the digital screening process before the UTI consultation. The first 3 digits of each patient’s zip code were used to determine geographic location.

#### Symptom presentation groups

Patients reported the presence of the following 4 uncomplicated UTI symptoms: urinary frequency, urinary urgency, dysuria (pain or burning while urinating), and hematuria (blood in the urine). Patients reported the presence of the following complicated UTI symptoms: an oral temperature greater than 99.5 °F; new onset of nausea and/or vomiting; new onset of feelings of general achiness/feeling unwell; back, abdominal, side, or groin pain; international travel within the past 6 months; and dehydration within the past 2 to 3 weeks [5,27,28]. Patients identified the presence of any vaginal infection symptoms: green, yellow, clear, or white discharge; odor from urine/discharge; and vaginal itching, blisters, lesions, or rash. Patients were grouped into 3 categories according to their reported symptom presentation: uncomplicated UTI symptoms, complicated UTI symptoms, and other (see Table 1). Appointments for recurrent infection (i.e., at least 2 appointments in 6 months, aside from relapse, or at least 3 appointments in 12 months, aside from relapse) were considered a complicated UTI, as long as the patient reported UTI symptoms (even if the patient had reported only symptoms of uncomplicated UTI during their recurrent visits) [19]. The presence or absence of vaginal symptoms was also assessed.

**Table 1.** Symptom groups and descriptions.

| Symptom Group | Description | N (%)<br>(N=51,474) |
| --- | --- | --- |
| Uncomplicated UTI<sup>1,3</sup> | Patients reporting $\geq 2$ of the 4 UTI symptoms, with or without symptoms of vaginal infection<br>Not a recurrent infection (first infection in 6 months, or $< 3$ infections in 12 months) | 31,730 (61.6) |
| Complicated UTI<sup>2,3</sup> | Patients who reported $\geq 1$ of the 4 UTI symptoms AND $\geq 1$ symptom of complicated infection, with or without symptoms of vaginal infection (regardless of recurrent infection)<br>OR<br>Any patient with recurrent infection ($\geq 2$ infections in 6 months or $\geq 3$ infections in 12 months) with or without symptoms of vaginal infection (regardless of having only uncomplicated UTI symptoms) | 18,788 (36.5) |
| Other<sup>3</sup> | Patients reporting $\leq 1$ of the 4 UTI symptoms, without symptoms of complicated infection, with or without symptoms of vaginal infection<br>Patients reporting none of the 4 UTI symptoms with $\leq 1$ of the complicated infection symptoms, with or without symptoms of vaginal infection<br>Regardless of recurrent infection | 956 (1.9) |

UTI, urinary tract infection.
<sup>1</sup>Uncomplicated UTI symptoms consisted of urinary frequency, urinary urgency, dysuria, and hematuria.
<sup>2</sup>Complicated UTI symptoms consisted of an oral temperature greater than 99.5 °F; new onset of nausea and/or vomiting; new onset of feelings of general achiness/not feeling well; back, abdominal, side, or groin pain; international travel within the past 6 months; and dehydration within the past 2–3 weeks.
<sup>3</sup>Symptoms of vaginal infections consisted of green, yellow, clear, or white discharge; odor from urine or discharge; and vaginal itching, blisters, lesions, or rash.

#### Prescription use and antibiotic type

Prescription use and antibiotic type were measured by analyzing the number of appointments that resulted in prescriptions and the type and number of medications that were prescribed. A standard clinical protocol was developed based on guidelines for uncomplicated UTIs; thus, the recommended antibiotics were nitrofurantoin, trimethoprim-sulfamethoxazole, and fosfomycin (see S1 Appendix) [18,22,26]. No standardized protocol was developed for complicated UTIs and/or symptoms of vaginal infection. Physicians were permitted to manage these cases according to their evaluation and discretion. However, specific population-guided protocols were developed (e.g., for women older than 65 years, patients with dual infections).

#### Symptom resolution

Patients received a survey 7 days after their appointment asking them to indicate whether their symptoms had resolved.

#### Treatment failure and relapse

Appointments that occurred within 30 days after an initial appointment were considered follow-up visits and reported as treatment failure or relapse. These visits were excluded from the original analyses and were instead reported as outcomes of the initial appointment. Due to insufficient data (e.g., it is unclear if most patients experienced any symptom resolution between appointments), treatment failure and relapse were reported together because these groups were indistinguishable.

### Statistical methods

Descriptive statistics were used to summarize the study sample, including frequencies, means, SDs, medians, and IQRs. Specifically, data were presented to examine prescription use and symptom resolution by age and US census region, as well as by grouping those with uncomplicated UTIs, complicated UTIs, and symptoms of vaginal infection. Number of prescriptions received, type of prescription, symptom resolution, and number of treatment failures/relapses by symptom profile category were also assessed. Symptom resolution rates were compared, using chi-square tests, between the complicated and uncomplicated UTI symptom groups and between all women who received a prescription medication with symptom resolution among 8 comparator studies, for a total of 9 tests [14,29-35]. Bonferroni correction was used to account for multiple tests, and p-values <0.005 were considered significant. All descriptive statistics and chi-square tests were calculated in R (version 4.0.5) for Macintosh.

## Results

The analysis set (n=51,474) included data from appointments with patients between the ages of 18 and 84 years (mean=39.4, SD=14.7). Most patients were between the ages of 22 to 46 years (n=30,423, 59.1%), and the South census geographic region had the greatest number of patients (n=20,172, 39.2%; see Table 2).

**Table 2.** Patient characteristics.^1^.

| Characteristics | N (%)<br>(N=51,474) |
| --- | --- |
| Age (years) |  |
| 18–21 | 3,791 (7.4) |
| 22–45 | 30,423 (59.1) |
| 46–65 | 14,449 (28.1) |
| ≥66 | 2,811 (5.5) |
| Region (US Census Region)<sup>2</sup> |  |
| South | 20,172 (39.2) |
| West | 17,851 (34.7) |
| Midwest | 9,970 (19.4) |
| Northeast | 3,481 (6.8) |

<sup>1</sup>Raw data counts by age and geographic region for each appointment. Individuals who used the program multiple times appear more than once in this table.
<sup>2</sup>Regions were determined via the US Census regions. West: Alaska, Arizona, California, Colorado, Hawaii, Idaho, Montana, Nevada, New Mexico, Oregon, Utah, Washington, Wyoming. Northeast: Connecticut, Maine, Massachusetts, New Hampshire, New Jersey, New York, Pennsylvania, Rhode Island, Vermont. Midwest: Illinois, Indiana, Iowa, Kansas, Michigan, Minnesota, Missouri, Nebraska, North Dakota, Ohio, South Dakota, Wisconsin. South: Alabama, Arkansas, Delaware, District of Columbia, Florida, Georgia, Kentucky, Louisiana, Maryland, Mississippi, North Carolina, Oklahoma, South Carolina, Tennessee, Texas, Virginia, West Virginia.

### Symptom presentation

Overall, symptom prevalence was high for urinary frequency (n=48,574, 94.4%), urinary urgency (n=48,662, 94.5%), and dysuria (n=50,244, 97.6%), while fewer patients reported hematuria (n=8,381, 16.3%). The majority of appointments (n=31,730, 61.6%) consisted of patients with uncomplicated UTI symptoms, but a substantial number (n=18,788, 36.5%) reported complicated UTI symptoms (see Table 1). The remaining appointments (n=956, 1.9%) consisted of patients in the other symptoms group. In total, 9,894 (19.2%) reported at least 1 symptom of vaginal infection: 5,142 (16.2%) in the uncomplicated UTI symptoms group, 4,618 (24.6%) in the complicated UTI symptoms group, and 134 (14.0%) in the other symptoms group.

### Prescription use and antibiotic choice

Patients from 50,826 (98.7%) appointments received at least 1 prescription, with 2,818 (5.5%) receiving more than 1 prescription. Patients from 44,304 (86.1%) appointments received 1 antibiotic, 2,320 (4.5%) received at least 1 antibiotic and an additional medication (e.g., phenazopyridine or fluconazole), 498 (1.0%) received 2 or more antibiotics without an additional medication, and 5 (<0.01%) received 1 or more other medications without an antibiotic. Prescription data were unavailable for 3,966 (7.7%) patient appointments on the platform.

Overall, clinical protocol adherence to recommended prescribing patterns was high (n=43,932, 94.0%), and patients were most likely to receive prescriptions for antibiotics that were recommended in the clinical protocol (see Table 3). This pattern was consistent across patient groups: uncomplicated UTI symptoms (n=27,128, 94.4%), complicated UTI symptoms (n=16,070, 93.7%), and other symptoms group (n=772, 88.0%). Most women who reported at least 1 vaginal symptom, in addition to UTI symptoms, received a recommended antibiotic (n=8,396, 84.9%).

**Table 3.** Symptom groups and prescription counts.

| Symptom Group | Received a Prescription<sup>1</sup><br>N (%) | Unknown Prescription<sup>2</sup><br>N (%) | Received Antibiotic<br>N (%)<sup>3</sup> | Received “Recommended” Antibiotic<br>N (%)<sup>4</sup> | Nitrofurantoin<br>N (%)<sup>5</sup> | TMP-SMX<br>N (%)<sup>5</sup> | Fosfomycin<br>N (%)<sup>5</sup> | Ciprofloxacin (Regular or XR)<br>N (%)<sup>5, 6</sup> |
| --- | --- | --- | --- | --- | --- | --- | --- | --- |
| Total<br>N=51,474<br>(100%) | 50,826 (98.7) | 3,699 (7.2) | 46,754 (92.0) | 43,932 (94.0) | 24,581 (52.6) | 19,404 (41.5) | 23 (0.0) | 2,431 (5.2) |
| UTI<br>N=50,518<br>(98.1%) | 49,903 (98.8) | 3,657 (7.2) | 45,877 (91.9) | 43,160 (94.1) | 24,200 (52.7) | 19,010 (41.4) | 23 (0.0) | 2,346 (5.1) |
| Uncomplicated<br>N=31,730<br>(62.8%) | 31,431 (99.1) | 2,471 (7.8) | 28,727 (91.4) | 27,128 (94.4) | 15,235 (53.0) | 11,913 (41.5) | 15 (0.0) | 1,402 (4.9) |
| Complicated<br>N=18,788<br>(37.2%) | 18,472 (98.3) | 1,186 (6.3) | 17,150 (94.8) | 16,070 (93.7) | 8,965 (52.3) | 7,097 (41.4) | 8 (0.0) | 944 (5.5) |
| Other<br>N=965<br>(1.9%) | 923 (95.6) | 42 (4.4) | 877 (95.0) | 772 (88.0) | 381 (43.4) | 394 (44.9) | 0 (0.0) | 85 (9.7) |

TMP-SMX, trimethoprim-sulfamethoxazole; UTI, urinary tract infection; XR, extended-release.
<sup>1</sup>Patients who received >1 prescription are only included once in this column.
<sup>2</sup>Some patients received a prescription, but there was no information about the prescription available in the dataset.
<sup>3</sup>Percentage represents the number of patients who received ≥1 antibiotics out of the total number who received a prescription. Patients who received >1 type of antibiotic are only included once.
<sup>4</sup>A recommended antibiotic is defined by the clinical protocol, not by the patient’s self-reported symptoms, and include nitrofurantoin, TMP-SMX, and fosfomycin. The percentage represents the number of patients who received a recommended antibiotic out of the total
number who received an antibiotic prescription. Note that 76 people received a prescription for nitrofurantoin and TMP-SMX; they are only listed once in this column.
<sup>5</sup>The percentages represent the number of patients who received that prescription out of the number of patients who received an antibiotic.
<sup>6</sup>Ciprofloxacin was an alternative regimen in the clinical protocol.

### Symptom resolution

Symptom resolution data from the 7-day follow-up survey were available for 3,541 women, including 2,193 women in the uncomplicated UTI group, 1,241 women in the complicated UTI group, and 107 women in the other symptoms group. Across all groups, 99.4% of patients (n=3,521) received a prescription, and the majority of these patients reported 7-day symptom resolution (89.7%, n=3,160) compared with less than half of patients who did not receive a prescription (40.0%, n=8; see Table 4). The symptom resolution rate in the complicated UTI group was lower than that of the uncomplicated UTI group; however, this difference was not significant (see Fig 2). Among those with a UTI and at least one symptom of vaginal infection, the symptom resolution rates were similar for those with uncomplicated UTIs (86.2%) and complicated UTIs (85.5%). Across all groups, the symptom resolution rate among patients who received a prescription within this study was significantly higher than 4 of the 8 comparator studies (□^2^ = 20.0–374.1, p<0.001), significantly lower than 1 study (□^2^ =11.9, p<0.001), and not significantly different from the remaining 3 studies (see Fig 2).

**Table 4.**
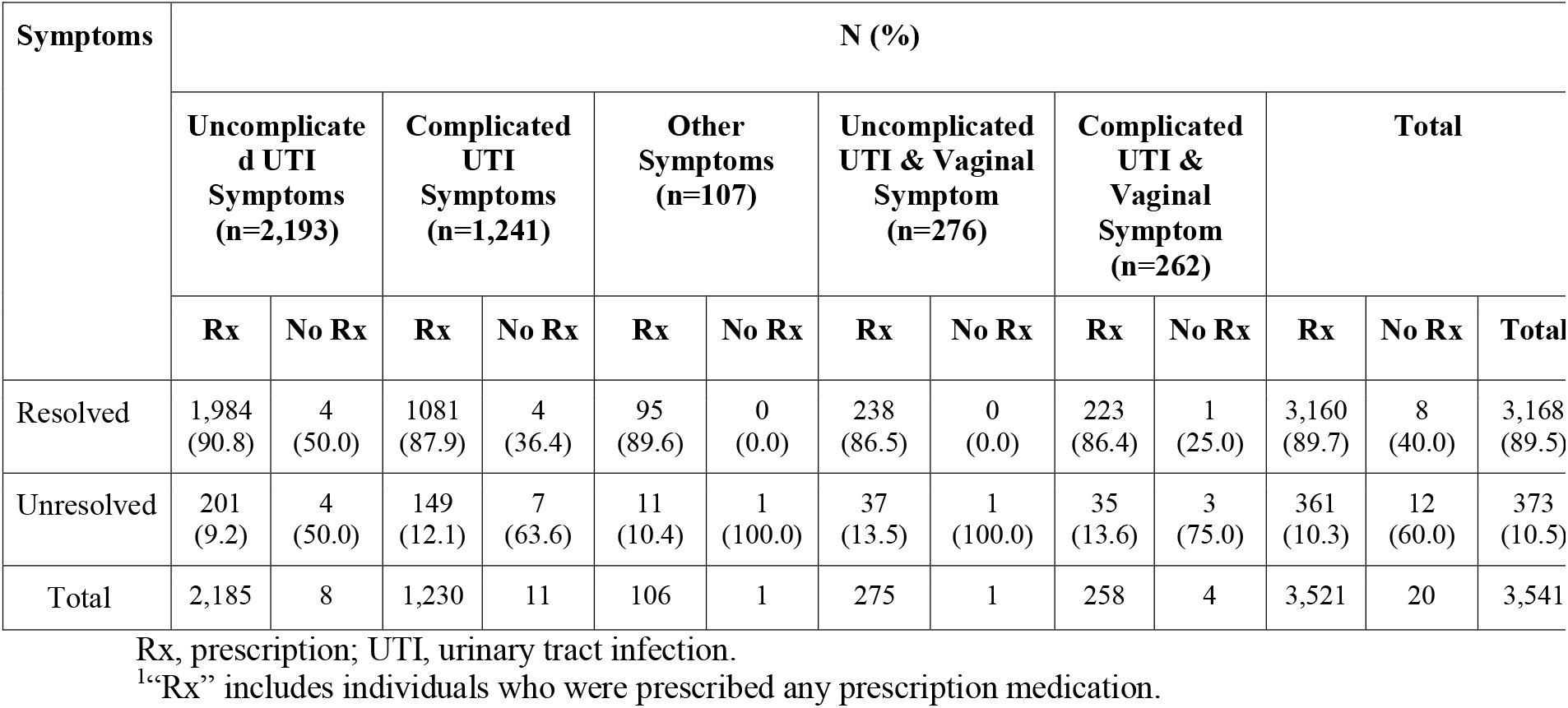
Symptom resolution stratified by symptom group and receiving a prescription or not.^1^.

| Symptoms | N (%) |  |  |  |  |  |  |  |  |  |  |  |  |
| --- | --- | --- | --- | --- | --- | --- | --- | --- | --- | --- | --- | --- | --- |
|  | Uncomplicated UTI Symptoms (n=2,193) |  | Complicated UTI Symptoms (n=1,241) |  | Other Symptoms (n=107) |  | Uncomplicated UTI & Vaginal Symptom (n=276) |  | Complicated UTI & Vaginal Symptom (n=262) |  | Total |  |  |
|  | Rx | No Rx | Rx | No Rx | Rx | No Rx | Rx | No Rx | Rx | No Rx | Rx | No Rx | Total |
| Resolved | 1,984 (90.8) | 4 (50.0) | 1081 (87.9) | 4 (36.4) | 95 (89.6) | 0 (0.0) | 238 (86.5) | 0 (0.0) | 223 (86.4) | 1 (25.0) | 3,160 (89.7) | 8 (40.0) | 3,168 (89.5) |
| Unresolved | 201 (9.2) | 4 (50.0) | 149 (12.1) | 7 (63.6) | 11 (10.4) | 1 (100.0) | 37 (13.5) | 1 (100.0) | 35 (13.6) | 3 (75.0) | 361 (10.3) | 12 (60.0) | 373 (10.5) |
| Total | 2,185 | 8 | 1,230 | 11 | 106 | 1 | 275 | 1 | 258 | 4 | 3,521 | 20 | 3,541 |

Rx, prescription; UTI, urinary tract infection.
<sup>1</sup>“Rx” includes individuals who were prescribed any prescription medication.

**Fig 2.**
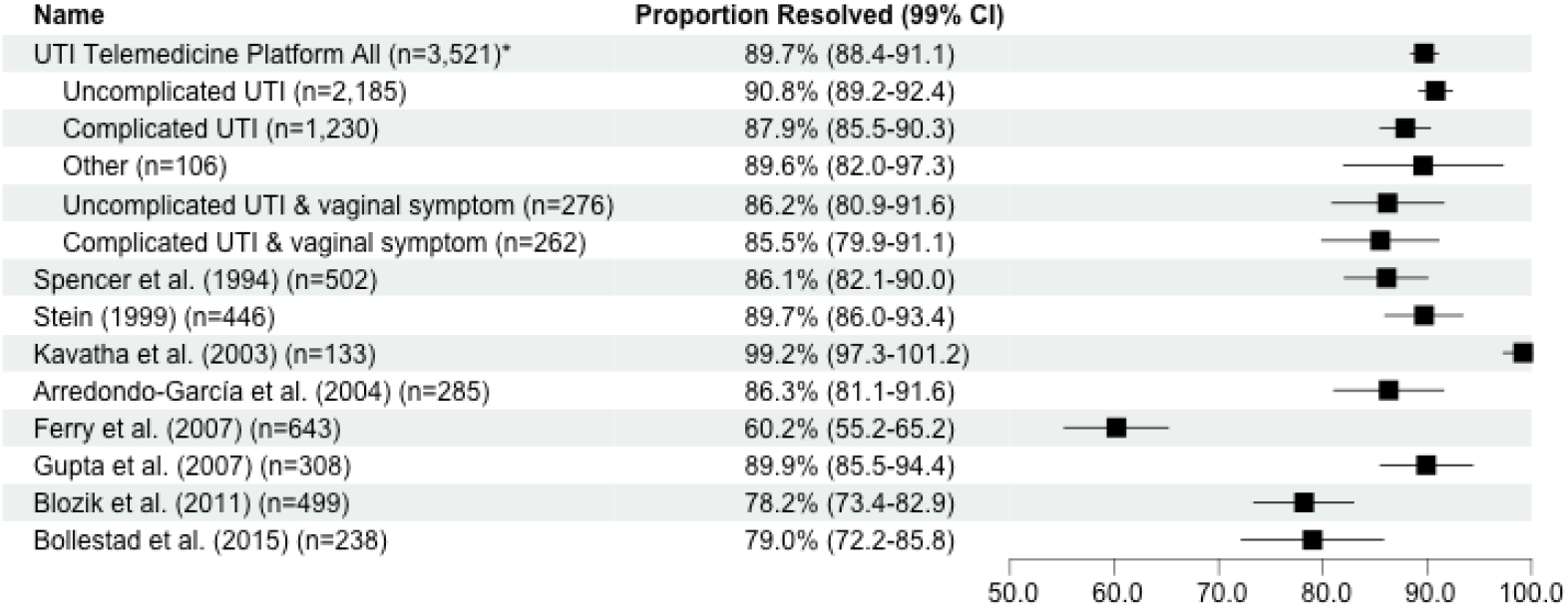
Forest plot of symptom resolution rates and 99% confidence intervals among all consultations, by UTI symptom group, and 8 previous studies’ reported rates. *”UTI Telemedicine Platform All” refers to all consultations with associated 7-day follow-up symptom resolution data from the current study, followed by each symptom category. Among the comparator studies, symptom resolution was measured on different days, including the following: 3 days [35], 4 days [14], 4 to 7 days [31], 5 to 9 days [32,34], 7 days [30], 8 to 10 days [33], and 9 to 15 days [29]. Of the 8 comparator studies, 7 used in-person standard-of-care [14,29-34] and 1 used telemedicine services [35]. If the comparator study was a clinical trial with a control group, then symptom resolution rates were only compared with treatment groups who received antibiotics [14].

### Treatment failure and relapse

Overall, 1,156 (2.2%) patients returned to use the program within 30 days after their original consultation: 264 patients (22.8%) returned within 7 days, 328 patients (28.4%) within 8 to 14 days, and 564 patients (48.8%) between 15 and 30 days after the initial appointment.

## Discussion

To the authors’ knowledge, this is the largest analysis of real-world data from a telemedicine service for treating UTIs to date. Data from 51,474 visits over 4 years indicated that patients from 98.1% of appointments self-reported at least 2 symptoms of uncomplicated or complicated UTIs, with dysuria being the most commonly reported symptom. More than half (52.4%) of the sample reported at least 1 symptom of complicated UTI or vaginal infection. Most appointments (98.7%) resulted in a prescription, and prescription use and type were similar regardless of patient-reported symptoms (uncomplicated UTI, complicated UTI, or vaginal infection). Most patients (94.0%) with available prescription data received antibiotics that were recommended in the protocol (nitrofurantoin, trimethoprim-sulfamethoxazole, and fosfomycin).

Of patients with available follow-up survey data, 89.5% reported symptom resolution, and resolution rates were similar between uncomplicated and complicated UTI groups. Furthermore, the symptom resolution rate across all UTI categories was similar to previous research on UTI telemedicine programs evaluating only uncomplicated UTI patients. Only 2.2% of patients returned to the program within 30 days after an initial appointment, suggesting low treatment failure and relapse rates. Resolution rates among uncomplicated and complicated UTIs were also similar to, or better than, previous reports of in-person standard of care (see Fig 2). Taken together, this analysis demonstrates the effectiveness of large-scale UTI telemedicine programs and extends those observations to include patients who self-report symptoms of complicated UTIs or vaginal infections. Although this study made no direct measurements regarding safety, a retrospective study evaluating the safety and efficacy of telemedicine management of uncomplicated UTIs in 526 women reported that teleprescription of antibiotics is as safe as prescriptions initiated during in-person consultations [35]. Overall, 78% of patients reported complete symptom resolution 3 days following teleconsultation, while 14% reported a decrease in uncomplicated UTI symptoms. Four percent reported deterioration, such as increased pain, flank pain, or fever, and 5% reported antibiotic adverse effects [35].

Previous research has reported that women perform well at self-diagnosing UTIs [36]. Despite self-reporting symptoms of complicated UTIs or vaginal infections, these data support the notion that the women using this telemedicine program were accurately able to self-identify that they had a UTI. Furthermore, through discussions with physicians, these patients may have clarified the presence and severity of complicated UTIs or vaginal symptoms, which may have emboldened physicians to provide care for women who self-reported these symptoms at intake. Women who self-reported complicated UTI and vaginal symptoms had similar resolution rates compared with those who reported uncomplicated UTIs symptoms. Thus, these data suggest that a telemedicine approach to identification and treatment of more complicated UTIs and vaginal symptoms may be effective. Further work to determine appropriate sub-grouping and standardized protocols for these conditions is likely warranted. Of note, if the program had excluded patients who reported complicated UTI or symptoms of vaginal infection, 24,064 women (approximately half of the sample) would have required in-person care, either through primary care, urgent care, or emergency department visits.

At scale, telemedicine programs that include uncomplicated and complicated UTIs could be a complementary offering at primary care clinics. This complementary service may result in reduced provider burdens within those settings, as well as within urgent care or emergency departments, freeing up valuable resources, without compromising quality of care or resolution rates. Undeniably, telemedicine access for UTI assessment and treatment can lessen the patient burden of in-person care. Future studies may identify the most effective protocols for integrating such services into the existing primary care setting, or even into urgent care centers and emergency departments.

Another noteworthy outcome of this analysis was that telemedicine physician adherence to recommended antibiotic protocols was higher than previously reported, according to concordance with the Infectious Diseases Society of America guidelines for antibiotics to treat UTIs [37,38]. The telemedicine physicians in this sample were adherent to these guidelines for 94.0% of appointments that resulted in antibiotic prescriptions. This high adherence rate may have been due to the implementation of the protocol and ease of use [39]. Moreover, provider prescribing practices were regularly audited to assure compliance. The recommended antibiotics in the clinical protocol were based on the guidelines for patients with uncomplicated UTIs [26,40]. Although not detailed in the current clinical protocol, the recommended antibiotic regimens may differ for patients exhibiting symptoms of pyelonephritis or complicated infection [40]. Telemedicine programs that allow for patients with symptoms of complicated UTIs or vaginal infections to receive appointments should include specific instructions for assessing the severity of the infection, the potential need for STI testing, and other medications that should be considered if a UTI diagnosis is not suspected.

### Limitations

As with all real-world evidence, the data used in this analysis were limited by what was collected through the clinical program. The present analysis relied on inferred diagnosis determined from digital responses and patient-reported symptoms at intake. Diagnoses made during treatment consultations were not obtained. As telemedicine programs become more ubiquitous, incorporating post-encounter diagnosis codes would provide greater granularity and understanding of treatment patterns and outcomes, particularly for those with more complicated cases or other symptoms that may be addressed as part of the telemedicine encounter. Integrating these types of programs into electronic health records diagnosis information would allow researchers to identify patients who inaccurately report their symptoms. Future work is needed to determine discrepancies between patient-reported symptoms on intake and physicians’ diagnoses based on direct patient encounters. This knowledge would help to expand clinical protocols to include guidance for determining the risk levels and appropriate next steps for patients who report symptoms of complicated UTI or vaginal infection.

The current analysis evaluated symptom resolution data for a total of 3,541 appointments, representing only 6.9% of the population. Although this still represents a large cohort in both the uncomplicated and complicated UTI groups, we can only speculate a similar rate of symptom resolution over the entire sample population. Notably, patients who did not experience symptom resolution were offered additional care, including referral to in-person care, and as such no symptom resolution data may have been available. A small percentage of patients (2.2%) returned to use the program within 30 days after the initial consultation, suggesting that most patients experienced a full recovery or sought in-person/other care, a feature this analysis cannot resolve. Patient follow-up data are needed to better understand symptom resolution when using direct-to-consumer telemedicine services. Future research may provide insight into how to ensure patients self-report symptom resolution, or perhaps, how data from electronic health records may be used to assess follow-up engagement with in-person care services.

## Conclusion

Telemedicine services make it easier and faster for patients to receive treatment for UTIs by eliminating travel and waiting time at emergency rooms. This analysis demonstrates that telemedicine programs are an effective and scalable option for treatment of UTIs, even for women who self-report symptoms of complicated UTIs or vaginal infection. Most patients in the program exhibited symptoms of uncomplicated or complicated UTIs. Of those patients with available follow-up data, most experienced symptom resolution within 7 days after their appointment. Implementing and scaling UTI telemedicine programs and including women who self-report symptoms of complicated UTI or vaginal infection can reduce the burden on urgent care facilities and emergency departments, without compromising on the quality of care.

## Data Availability

All data produced in the present study are available upon reasonable request to the authors

## Abbreviations

STIs: sexually transmitted infections
UTIs: urinary tract infections

## Acknowledgments

The authors would like to acknowledge the following people who developed and managed the telemedicine program evaluated in this study: Yvette Gaudreau, Lara Goorland, Anthony Dearman, Gabe Gaviola, Adina Schwartz, Jennifer Jing, Emily Miller, and Doug Elwood. Additionally, writing support was provided by Virgo Health.

## S1 Appendix: Prescription Information

| **Recommended Regimens** |  |  |
| --- | --- | --- |
| Nitrofurantoin<br>100 mg, twice per day for 5 days | Trimethoprim-sulfamethoxazole<br>160–800 mg twice per day for 3 days | Fosfomycin<br>3 g single dose |
| **Alternative Regimens** |  |  |
| Ciprofloxacin<sup>*</sup><br>250 mg twice per day for 3 days | Ciprofloxacin extended release<sup>*</sup><br>500 mg once per day for 3 days | Levofloxacin<sup>*</sup><br>250 mg once per day for 3 days |
| Ofloxacin<sup>*</sup><br>400 mg single dose | Amoxicillin-clavulanate<sup>†</sup><br>500–125 mg twice per day for 3–7 days | Cefpodoxime<sup>†</sup><br>100 mg twice per day for 3–7 days |
| Cefdinir<sup>†</sup><br>300 mg twice per day for 3–7 days | Cefaclor<sup>†</sup><br>250 mg or 500 mg 3 times per day for 3–7 days | Cefadroxil<sup>†</sup><br>500 mg twice per day for 3–7 days |

<sup>\*</sup>Fluoroquinolones
<sup>†</sup>Beta lactams

## References

1. Schappert SM, Rechtsteiner EA. Ambulatory medical care utilization estimates for 2007. Vital Health Stat 13. 2011;(169):1–38.

2. Chou SC, Baker O, Schuur JD. Changes in emergency department care intensity from 2007-16: analysis of the National Hospital Ambulatory Medical Care survey. West J Emerg Med. 2020;21(2):209–216. doi:10.5811/westjem.2019.10.43497.

3. Stein JC, Navab B, Frazee B, Tebb K, Hendey G, Maselli J, et al. A randomized trial of computer kiosk-expedited management of cystitis in the emergency department. Acad Emerg Med. 2011;18(10):1053–1059. doi:10.1111/j.1553-2712.2011.01167.x.

4. Bashshur RL, Reardon TG, Shannon GW. Telemedicine: a new health care delivery system. Annu Rev Public Health. 2000;21:613–637. doi:10.1146/annurev.publhealth.21.1.613.

5. Bono MJ, Leslie SW, Reygaert WC. Urinary tract infection. In: StatPearls [Internet]. Treasure Island (FL): StatPearls Publishing; 2022.

6. Taur Y, Smith MA. Adherence to the Infectious Diseases Society of America guidelines in the treatment of uncomplicated urinary tract infection. Clin Infect Dis. 2007;44(6):769–774. doi:10.1086/511866.

7. Foxman B. Urinary tract infection syndromes: occurrence, recurrence, bacteriology, risk factors, and disease burden. Infect Dis Clin North Am. 2014;28(1):1–13. doi:10.1016/j.idc.2013.09.003.

8. Ozturk T, Talo M, Yildirim EA, Baloglu UB, Yildirim O, Acharya UR. Automated detection of COVID-19 cases using deep neural networks with X-ray images. Comput Biol Med. 2020;121:103792. doi:10.1016/j.compbiomed.2020.103792.

9. Campbell J, Felver M, Kamarei S. ‘Telephone treatment’ of uncomplicated acute cystitis. Cleve Clin J Med. 1999;66(8):495–501. doi:10.3949/ccjm.66.8.495.

10. Saint S, Scholes D, Fihn SD, Farrell RG, Stamm WE. The effectiveness of a clinical practice guideline for the management of presumed uncomplicated urinary tract infection in women. Am J Med. 1999;106(6):636–641. doi:10.1016/s0002-9343(99)00122-9.

11. Barry HC, Hickner J, Ebell MH, Ettenhofer T. A randomized controlled trial of telephone management of suspected urinary tract infections in women. J Fam Pract. 2001;50(7):589–594.

12. Vinson DR, Quesenberry CP, Jr. The safety of telephone management of presumed cystitis in women. Arch Intern Med. 2004;164(9):1026–1029. doi:10.1001/archinte.164.9.1026.

13. Schauberger CW, Merkitch KW, Prell AM. Acute cystitis in women: experience with a telephone-based algorithm. WMJ. 2007;106(6):326–329.

14. Bollestad M, Grude N, Lindbaek M. A randomized controlled trial of a diagnostic algorithm for symptoms of uncomplicated cystitis at an out-of-hours service. Scand J Prim Health Care. 2015;33(2):57–64. doi:10.3109/02813432.2015.1041827.

15. Rastogi R, Martinez KA, Gupta N, Rood M, Rothberg MB. Management of urinary tract infections in direct to consumer telemedicine. J Gen Intern Med. 2020;35(3):643–648. doi:10.1007/s11606-019-05415-7.

16. Bent S, Nallamothu BK, Simel DL, Fihn SD, Saint S. Does this woman have an acute uncomplicated urinary tract infection? JAMA. 2002;287(20):2701–2710. doi:10.1001/jama.287.20.2701.

17. Little P, Moore MV, Turner S, Rumsby K, Warner G, Lowes JA, et al. Effectiveness of five different approaches in management of urinary tract infection: randomised controlled trial. BMJ. 2010;340:c199. doi:10.1136/bmj.c199.

18. Gupta K, Grigoryan L, Trautner B. Urinary tract infection. Ann Intern Med. 2017;167(7):ITC49–ITC64. doi:10.7326/aitc201710030.

19. Sabih A, Leslie SW. Complicated urinary tract infections. 2022 May 27. In: StatPearls [Internet]. Treasure Island (FL): StatPearls Publishing; 2022.

20. Smith WB, I., Kohlwes RJ. From leather bags to webcams, the emerging tools of tele-primary care. J Gen Intern Med. 2020;35(3):628–629. doi:10.1007/s11606-019-05603-5.

21. Giesen LGM, Cousins G, Dimitrov BD, van de Laar FA, Fahey T. Predicting acute uncomplicated urinary tract infection in women: a systematic review of the diagnostic accuracy of symptoms and signs. BMC Fam Pract. 2010;11:78. doi:10.1186/1471-2296-11-78.

22. Colgan R, Williams M. Diagnosis and treatment of acute uncomplicated cystitis. Am Fam Physician. 2011;84(7):771–776.

23. Garland SM, Tabrizi SN. Diagnosis of sexually transmitted infections (STI) using self-collected non-invasive specimens. Sex Health. 2004;1(2):121–126. doi:10.1071/sh03014.

24. Alidjanov JF, Naber KG, Abdufattaev UA, Pilatz A, Wagenlehner FM. Reliability of symptom-based diagnosis of uncomplicated cystitis. Urol Int. 2019;102(1):83–95. doi:10.1159/000493509.

25. Goebel MC, Trautner BW, Grigoryan L. The five Ds of outpatient antibiotic stewardship for urinary tract infections. Clin Microbiol Rev. 2021;34(4):e0000320. doi:10.1128/cmr.00003-20.

26. Grigoryan L, Trautner BW, Gupta K. Diagnosis and management of urinary tract infections in the outpatient setting: a review. JAMA. 2014;312(16):1677–1684. doi:10.1001/jama.2014.12842.

27. U.S. Department of Health and Human Services. Complicated urinary tract infections: developing drugs for treatment. Guidance for industry. 2018. Available at https://www.fda.gov/regulatory-information/search-fda-guidance-documents/complicated-urinary-tract-infections-developing-drugs-treatment. Accessed on April 20, 2022.

28. Belyayeva M, Jeong JM. Acute pyelonephritis. 2022 Jul 5. In: StatPearls [Internet]. Treasure Island (FL): StatPearls Publishing; 2022.

29. Spencer RC, Moseley DJ, Greensmith MJ. Nitrofurantoin modified release versus trimethoprim or co-trimoxazole in the treatment of uncomplicated urinary tract infection in general practice. J Antimicrob Chemother. 1994;33 Suppl A:121–129. doi:10.1093/jac/33.suppl_a.121.

30. Stein GE. Comparison of single-dose fosfomycin and a 7-day course of nitrofurantoin in female patients with uncomplicated urinary tract infection. Clin Ther. 1999;21(11):1864–1872. doi:10.1016/s0149-2918(00)86734-x.

31. Kavatha D, Giamarellou H, Alexiou Z, Vlachogiannis N, Pentea S, Gozadinos T, et al. Cefpodoxime-proxetil versus trimethoprim-sulfamethoxazole for short-term therapy of uncomplicated acute cystitis in women. Antimicrob Agents Chemother. 2003;47(3):897–900. doi:10.1128/aac.47.3.897-900.2003.

32. Arredondo-García JL, Figueroa-Damián R, Rosas A, Jáuregui A, Corral M, Costa A, et al. Comparison of short-term treatment regimen of ciprofloxacin versus long-term treatment regimens of trimethoprim/sulfamethoxazole or norfloxacin for uncomplicated lower urinary tract infections: a randomized, multicentre, open-label, prospective study. J Antimicrob Chemother. 2004;54(4):840–843. doi:10.1093/jac/dkh414.

33. Ferry SA, Holm SE, Stenlund H, Lundholm R, Monsen TJ. Clinical and bacteriological outcome of different doses and duration of pivmecillinam compared with placebo therapy of uncomplicated lower urinary tract infection in women: the LUTIW project. Scand J Prim Health Care. 2007;25(1):49–57. doi:10.1080/02813430601183074.

34. Gupta K, Hooton TM, Roberts PL, Stamm WE. Short-course nitrofurantoin for the treatment of acute uncomplicated cystitis in women. Arch Intern Med. 2007;167(20):2207–2212. doi:10.1001/archinte.167.20.2207.

35. Blozik E, Sommer-Meyer C, Cerezo M, von Overbeck J. Effectiveness and safety of telemedical management in uncomplicated urinary tract infections. 2011;17(2):78–82. doi:10.1258/jtt.2010.100406.

36. Gupta K, Hooton TM, Roberts PL, Stamm WE. Patient-initiated treatment of uncomplicated recurrent urinary tract infections in young women. Ann Intern Med. 2001;135(1):9–16. doi:10.7326/0003-4819-135-1-200107030-00004.

37. Zatorski C, Zocchi M, Cosgrove SE, Rand C, Brooks G, May L. A single center observational study on emergency department clinician non-adherence to clinical practice guidelines for treatment of uncomplicated urinary tract infections. BMC Infect Dis. 2016;16(1):638. doi:10.1186/s12879-016-1972-6.

38. Grigoryan L, Zoorob R, Wang H, Trautner BW. Low concordance with guidelines for treatment of acute cystitis in primary care. Open Forum Infect Dis. 2015;2(4):ofv159. doi:10.1093/ofid/ofv159.

39. Grigoryan L, Nash S, Zoorob R, Germanos GJ, Horsfield MS, Khan FM, et al. Qualitative analysis of primary care provider prescribing decisions for urinary tract infections. Antibiotics (Basel). 2019;8(2). doi:10.3390/antibiotics8020084.

40. Gupta K, Hooton TM, Naber KG, Wult B, Colgan R, Miller LG, et al. International clinical practice guidelines for the treatment of acute uncomplicated cystitis and pyelonephritis in women: a 2010 update by the Infectious Diseases Society of America and the European Society for Microbiology and Infectious Diseases. Clin Infect Dis. 2011;52(5):e103–e120. doi:10.1093/cid/ciq257.

